# Human gain-of-function variants in HNF1A confer protection from diabetes but independently increase hepatic secretion of multiple cardiovascular disease risk factors

**DOI:** 10.1101/2022.03.29.22273133

**Authors:** Natalie DeForest, Babu Kavitha, Siqi Hu, Roi Isaac, Minxian Wang, Xiaomi Du, Camila De Arruda Saldanha, Jenny Gylys, Ruben Abagyan, Laeya Najmi, Viswanathan Mohan, Alnylam Human Genetics, AMP-T2D Consortium, Jason Flannick, Gina M. Peloso, Philip Gordts, Sven Heinz, Aimee M. Deaton, Amit V. Khera, Jerrold Olefsky, Venkatesan Radha, Amit R. Majithia

## Abstract

Loss-of-function mutations in Hepatocyte Nuclear Factor 1A (HNF1A) are known to cause rare forms of diabetes and alter hepatic physiology through unclear mechanisms. In the general population, 1:100 individuals carry a rare protein-coding variant in *HNF1A*, most of unknown functional consequence. To characterize the full allelic series, we performed deep mutational scanning of 11,970 protein-coding *HNF1A* variants in human hepatocytes and clinical correlation with 553,246 exome-sequenced individuals. Surprisingly, we found ∼1:5 rare protein-coding *HNF1A* variants found in the general population cause molecular gain-of-function (GOF), increasing the transcriptional activity of HNF1A by up to 50%. GOF in HNF1A conferred protection from type 2 diabetes (T2D) (OR=0.60, p=8.4 x 10^-7^), but not against coronary artery disease. Independently of T2D, increased hepatic expression of *HNF1A* promoted a pro-inflammatory and pro-atherogenic serum profile mediated in part by enhanced transcription of risk genes including *PCSK9*. In summary, ∼1:300 individuals carry a GOF variant in *HNF1A* that protects carriers from diabetes but enhances hepatic secretion of metabolic disease risk factors.

---

Hepatocyte Nuclear Factor 1 alpha (*HNF1A*) is a lineage-determining transcription factor expressed in several tissues including the liver and pancreas^1^. Rare, complete loss-of-function mutations (LOF) in *HNF1A* have been shown to cause an autosomal dominant monogenic form of diabetes (MODY3)^2, 3^ through deficiencies in pancreatic insulin secretion via a haploinsufficient genetic mechanism^4^. MODY3 patients have been observed to carry altered levels of serum lipoproteins^5^ and lower levels of the inflammatory marker high-sensitivity C-reactive protein (hsCRP)^6^ than matched patients with type 2 diabetes (T2D), suggesting a potential liver-mediated functional role for HNF1A in metabolic disease pathogenesis. However, diabetes itself through underlying insulin resistance or elevated blood glucose can dysregulate serum lipids^7^ and systemic inflammation^8^, and thus these observations in diabetic individuals with MODY3 cannot evidentiate HNF1A as a direct hepatic regulator of serum lipids or inflammation. Population-based genetic association studies have corroborated associations between *HNF1A* and T2D^9^, serum lipoproteins^10^, and hsCRP^11^, but do not provide insight into mechanism as most of the associated variants (SNPs) are either non-coding or have minimal experimentally observable consequences on protein function^12^. Finally, common HNF1A SNPs have been associated with coronary artery disease (CAD) risk^13,14^ but these SNPs are also associated with T2D, lipid levels and hsCRP which are known, independent CAD mediators and risk factors^15,16^. Thus, the relationship between HNF1A function, these risk factors, and the specific mechanisms of disease pathogenesis remains unclear.

Characterization of an allelic series of functional variants for the hepatic functions of *HNF1A* with clinical correlation would enable a mechanistic understanding of the complex role for *HNF1A* in driving metabolic disease through risk factors in multiple tissues including the pancreas (via diabetes) and liver (inflammation/hsCRP and serum lipids) with their respective directions-of-effect. In the past few years, dozens of novel protein-coding alleles in *HNF1A* have been identified from population-based exome sequencing^17^. These variants are individually rare (minor allele frequency (MAF) < 0.001) and mostly of unknown functional consequence, but in aggregate are carried by over one percent of the population.

To address this gap in knowledge, we examined the full allelic series of protein-coding variants in *HNF1A* using deep mutational scanning, a high-throughput approach that has been successfully utilized to characterize protein-coding variants in clinically important genes^18–20^. All possible single amino acid substitutions in HNF1A were tested for transcriptional activity in human hepatocytes. These comprehensive data were intersected with carriers of rare protein-coding variants (MAF<0.001) identified from among 553,246 sequenced individuals in order to relate variant to function to phenotype for metabolic disease risk factors and disease outcomes including T2D and CAD. We provide the first description of gain-of-function (GOF) variants which while conferring protection from T2D promote serum inflammation, thrombogenic gene expression, and an atherogenic lipid profile through liver specific enhancement of HNF1A target genes.

## Results

### Identification of gain-of-function substitutions from comprehensive functional testing of 11,970 HNF1A amino acid variants in human hepatocytes

To comprehensively assess the hepatic function of protein-coding variants in *HNF1A*, we synthesized a cDNA library comprised of all possible single amino acid (aa) permutations of HNF1A (630 aa protein x 19 aa changes = 11,970, Supplementary Table 1) such that each transgene encoded a single amino acid substitution. The library was introduced into a human hepatocyte cell line previously engineered to lack endogenous *HNF1A* and at a dilution maximizing the number of cells receiving only a single copy of *HNF1A* transgene. The *HNF1A* transgenes were induced for six days via a doxycycline-inducible promoter and cells were sorted via fluorescence-activated cell sorting (FACS) based on their protein expression level of TM4SF4, a cell surface protein shown to be a direct transcriptional target of HNF1A in pancreas and liver^21,22^ (Figure 1A). The population of cells bearing *HNF1A* transgenes was partitioned into two bins separated by a ten-fold expression intensity difference in TM4SF4: TM4SF4^low^ and TM4SF4^high^ (Figure 1A, Supplementary Figure 1). To recover and quantify the HNF1A variants in each TM4SF4 expression bin, *HNF1A* transgenes from the TM4SF4^low^ and TM4SF4^high^ cells were sequenced by targeted next-generation sequencing. Raw function scores were generated for each amino acid substitution at each site in HNF1A by determining the frequency of appearance of each variant in the TM4SF4^low^ and TM4SF4^high^ bins as we and others have previously done^19,23^. Over 99% of the originally designed variants were recovered in sufficient quantity from the pooled screen to be assigned a function score (Supplementary Figure 2 and Supplementary Table 2).

**Figure 1.**
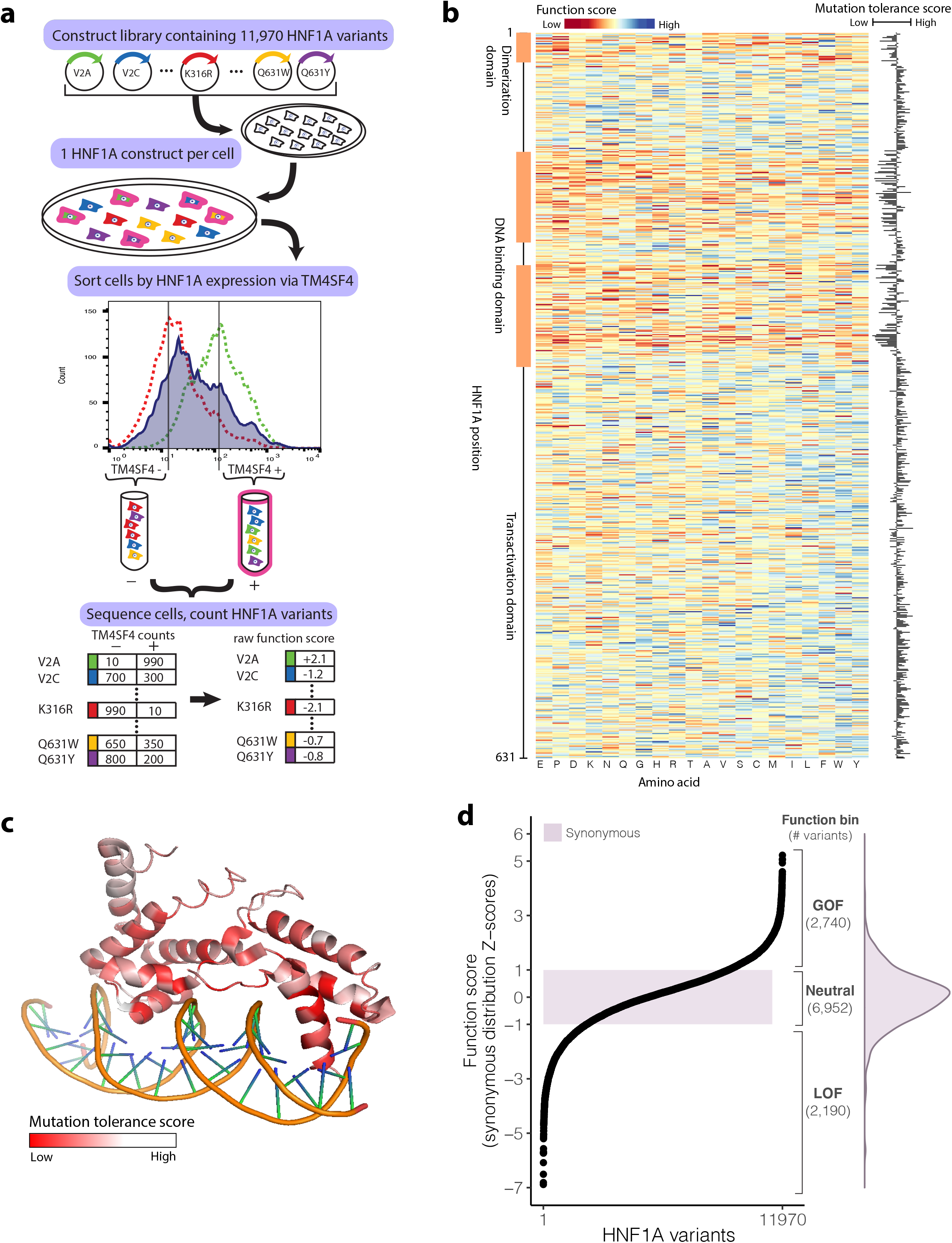
Identification of gain-of-function variants from comprehensive functional testing of 11,970 HNF1A variants in human hepatocytes. **a**) A library of 11,970 *HNF1A* constructs was synthesized, with each construct encoding a single amino acid substitution. The construct library was introduced into HUH7 hepatocytes (deleted for endogenous *HNF1A*) at a dilution of one construct per cell. The resulting polyclonal population of HUH7 hepatocytes was separated via FACS according to the expression of the known HNF1A transcriptional target TM4SF4 and sorted into low (−) and high (+) bins of HNF1A activity. Activity cutoffs were established through flow cytometry experiments of *HNF1A* KO cells (dashed red line) and WT cells (dashed green line). Each bin of cells was sequenced at the transgenic HNF1A locus to identify and tabulate the introduced variants. Each HNF1A variant was assigned a function score based on its abundance in the low and high TM4SF4 expression bins. **b**) Heatmap of 11,970 HNF1A variant function scores, arranged according to the primary amino acid sequence (rows). Function scores lower than WT are shaded red and function scores greater than WT are shaded blue. Function scores summed by amino acid position are plotted to the right, showing the level of tolerance for any amino acid substitution away from WT at each position. **c**) Mutation tolerance scores as described in **b**) overlaid on the crystalized protein structure of HNF1A DNA-binding domain (PF04814). Positions intolerant of amino acid changes (i.e. lower function scores) are shaded red. Helices that make direct contacts with the DNA are the most intolerant of mutations. **d**) (left panel) HNF1A function scores ranked for all 11,970 amino acid variants tested. The shaded purple box represents +/- 1 Z-score of the distribution of 613 synonymous HNF1A variants tested with the corresponding distribution (right panel). Function bins correspond to variants with function scores above (GOF), within (neutral), or below (LOF) +/- 1 Z-score of the synonymous distribution, shown with the total number of variants per bin.

The raw function scores for all protein-coding *HNF1A* variants were compared with the known sequence:function relationships of HNF1A. First, the function scores were averaged at each of the 631 amino acid positions along HNF1A to obtain a “mutation tolerance” score representing the effect on function of substituting any non-WT amino acid at that position; i.e. low “mutation tolerance” represents a functionally conserved residue (Figure 1B). Several clusters of amino acids along the primary sequence exhibiting low mutation tolerance scores were observed and colocalized with known domains critical for HNF1A molecular function including the dimerization domain and the DNA binding domain^24,25^. The mutation tolerance scores were also overlaid onto the HNF1A DNA binding domain crystal structure to evaluate the relationship between function and higher-order structure (Figure 1C). Within the extent of the available crystal structure, the amino acid residues comprising the alpha helices in the DNA binding domain that directly contact the DNA double helix exhibit the lowest mutation tolerance scores, consistent with a high degree of functional conservation (Figure 1C).

Taking a conservative approach to minimize the effect of extreme values, the raw individual variant scores were added to the mutation tolerance score at that amino acid position (Figure 1B) and rescaled to the mean and standard deviation (SD) of the distribution of synonymous variants to provide an intuitive interpretation to the function scores (Figure 1D). Of the 11,970 variants tested, 2,190 fell into the putative LOF category (score < -1SD), 6,952 fell into the neutral category (-1SD <= score <= 1SD) and surprisingly 2,740 fell into a putative gain-of-function GOF (score > 1SD) category indicating that those amino acid substitutions increased the transcriptional activity of HNF1A over WT in our assay, a finding not previously reported in over two decades of HNF1A functional variant characterization^2^. As expected, variants categorized as putative LOF by our experiments occurred at codons that were more evolutionarily conserved in mammalian genomic sequences^26^ than those classified as neutral (p < 10^-6^ Wilcoxon rank-sum test; Supplementary Figure 3). Variants categorized as putative GOF, however, could not be distinguished from neutral variants on the basis of evolutionary conservation suggesting that they would be poorly annotated by computational prediction algorithms leveraging conservation metrics^27,28^. To explicitly quantify on a per variant basis the confidence of LOF/GOF, a posterior error probability^29^ (PEP) was computed for every variant based on where its function score fell in the distribution of function scores for synonymous versus nonsynonymous variants (Figure 1D, Supplementary Table 2).

### Gain-of-function *HNF1A* variants are carried in the general population and increase transcriptional activity in multiple cellular contexts

To evaluate the human relevance of our deep mutational scan and assess if these putative GOF variants are present in people, we intersected our comprehensive dataset of HNF1A function scores with *HNF1A* protein-coding variants in the UK Biobank, a population-based cohort containing 454,756 exome sequenced individuals^30,31^. From the exome sequences we extracted all *HNF1A* variants and identified 4,302 individuals who carried a protein-coding variant with minor allele frequency less than 0.001. Of the 444 unique protein-coding variants identified, 369 (83%) were absent from existing archives of clinically relevant variants (ClinVar^32^; Supplementary Table 3). The distribution of HNF1A function scores in the UK Biobank peaked at the same level as the distribution of synonymous variants (Figure 2A), consistent with prior observations that most protein-coding variants are not functional^33^. The distribution of function scores in the UK Biobank also notably lacked the lowest scoring pathogenic/MODY3 variants as would be expected for a generally healthy population-based cohort^30^ and with the estimated UK prevalence of MODY3 being ∼5 per 100,000^34^ (Supplementary Figure 4). The presence of LOF *HNF1A* variants in population-based cohorts has been described previously^35^ but remarkably carriers of gain-of-function (GOF) variants have not previously been described in the many studies functionally characterizing human genetic variants in *HNF1A*.

**Figure 2.**
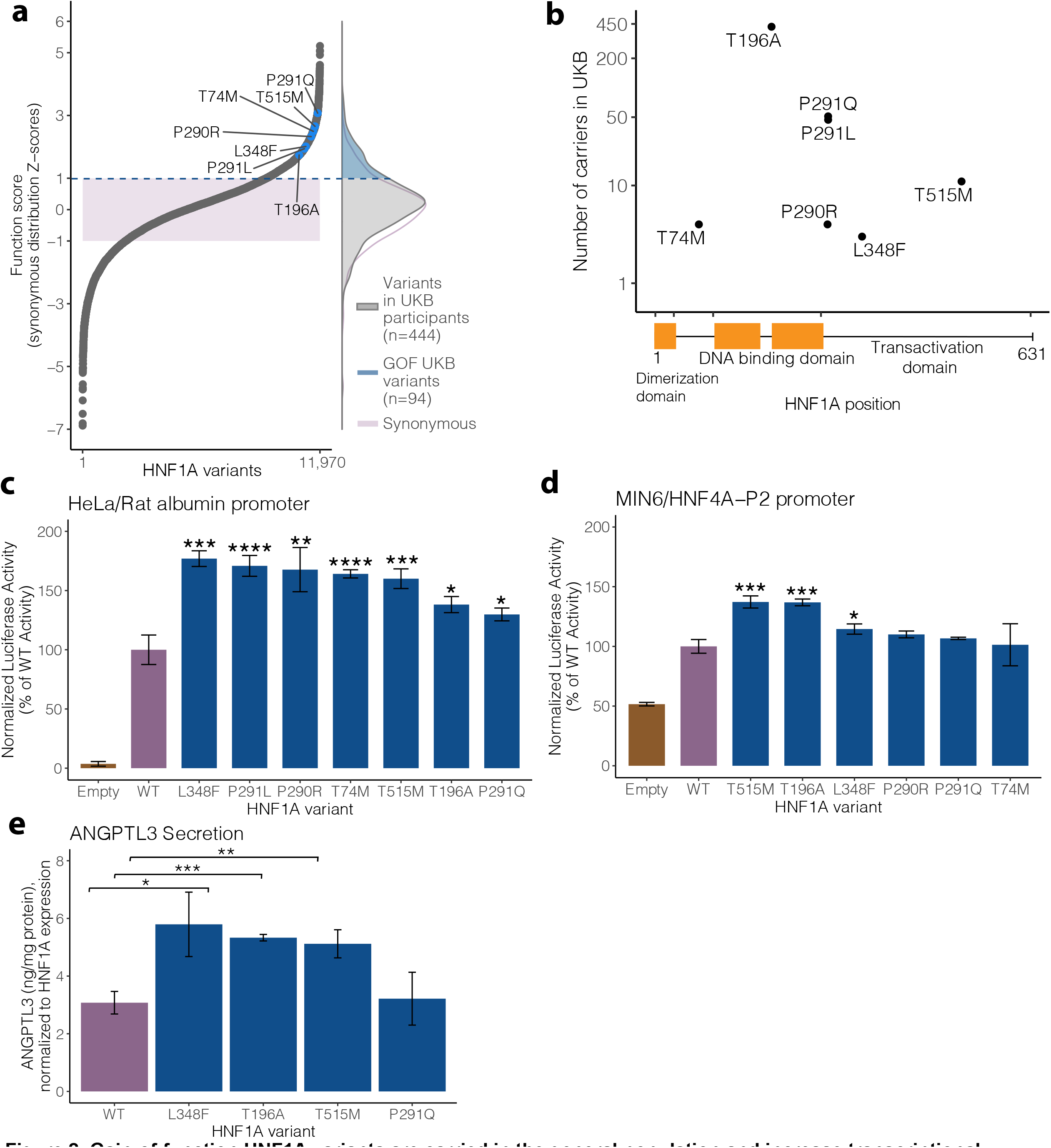
Gain-of-function HNF1A variants are carried in the general population and increase transcriptional activity in multiple cellular contexts. **a)** (left panel) Selected human HNF1A variants with function scores greater than WT highlighted in blue among the full distribution of all 11,970 amino acid variants tested. The shaded purple box represents +/- 1 Z-score of the distribution of 613 synonymous HNF1A variants tested. (right panel) The distributions of function scores of rare protein coding *HNF1A* variants (n=444) identified from 454,756 sequenced individuals in the UK Biobank (UKB) and of the 613 synonymous variants. An upper tail of function increasing variants in UK Biobank individuals is highlighted in blue (n=94 unique variants in 1,469 individuals). **b**) Location of top scoring variants selected for validation along the HNF1A protein primary amino acid sequence with number of human variant carriers identified in 454,756 exome sequences from the UK Biobank. **c**) Putative GOF variants were individually recreated and tested for transcriptional activation using luciferase reporters in HeLa cells (which lack endogenous HNF1A activity) on the rat albumin promoter. Activity measurements are shown as percentages of WT HNF1A activity +/- s.e.m; n=3. Basal promoter activity in cells is measured by transfection of vectors lacking the HNF1A transgene (empty). (* p<0.05, ** p<0.01, *** p<0.001, **** p<0.0001, linear mixed model) **d**) The same variants described in b) were tested for transcriptional activity in mouse insulinoma MIN6 cells on the HNF4A-P2 promoter. e) GOF HNF1A variant constructs were transfected into HNF1A deleted HUH7 hepatocytes, and ANGPTL3 levels were measured by ELISA 48 hours post confluency (n=3 per variant). Transfection with HNF1A p.L348F, p.T196A, and p.T515M increases ANGPTL3 secretion as compared to WT (*** p<0.0005,** p<0.005, * p<0.05, linear mixed model).

To confirm and further characterize these putative GOF *HNF1A* variants carried by humans in the general population, we selected a series of high-scoring GOF variants that spanned across the protein domains of HNF1A (Figure 2A). These seven variants ranged in frequency from 3 carriers (p.L348F) to 421 carriers (p.T196A) in the 454,756 sampled exomes (Figure 2B). Each variant was recreated, transfected, and assessed for transcriptional activity using luciferase reporter assays as we^36^ and others^37^ have previously performed in two separate experimental contexts: 1) HeLa cells using the rat albumin promoter (Figure 2C) and 2) MIN6 insulinoma cells using the HNF4A-P2 promoter (Figure 2D). In HeLa cells, all selected variants showed significant increases in transcriptional activity over WT (ranging from 30-70%) whereas in MIN6 cells the measured increase in transactivation was attenuated with only three of the tested variants exceeding the nominal threshold of significance. Notably, the background activity of HNF1A in Hela cells was nil, whereas in MIN6 cells it approached fifty percent of WT. Given that GOF was observed in two independent HNF1A-free cellular contexts (i.e. HeLa and HUH7 *HNF1A*-null cells) the attenuation in signal in the MIN6 cell system likely reflected decreased sensitivity in measuring increased HNF1A transcription in the setting of pre-existing high levels of endogenous HNF1A activity.

To assess the functional consequence of GOF *HNF1A* in hepatocytes we developed a secretion assay for ANGPTL3, a hepatically secreted protein that regulates serum lipids^38^ whose gene is bound by HNF1A^39^ (Supplementary Figure 5). After confirming that HNF1A functionally regulates production of ANGPTL3 from cultured hepatocytes (Supplementary Figure 5b), we evaluated ANGPTL3 secretion in *HNF1A* null hepatocytes and performed reconstitution experiments with WT and the identified GOF *HNF1A* variants. The p.L348F, p.T196A, and p.T515M variants but not the p.P291Q increased ANGPTL3 secretion when transfected into *HNF1A*-null HUH7 hepatocytes compared to WT (Figure 2E). Taken together, these cell culture experiments demonstrate that GOF HNF1A variants increase transcriptional activity in both pancreatic and hepatic cellular contexts.

### *HNF1A* GOF variant carriers in the population are protected from T2D, but not CAD, and show elevated levels of serum inflammatory/atherogenic risk factors

We examined the relationship between HNF1A function and T2D in carriers of rare protein-coding *HNF1A* variants ascertained from the UK Biobank and found an inverse association between HNF1A function and T2D risk in the full multi-ancestry cohort (OR=0.60 per SD increase in HNF1A function, 95% CI=[0.49-0.73], p=7.8 x 10^-7^, n=4,302) (Figure 3A). A similar association was obtained when restricting to European ancestry *HNF1A* rare variant carriers (OR=0.58 per SD increase in HNF1A function, 95% CI=[0.46-0.74], p=5.6 x 10^-6^, n=3,089). Given the previously known associations between LOF variants in *HNF1A* and diabetes^2,35,40^, we performed a categorical analysis binning the *HNF1A* variants by function (GOF, neutral, LOF) and separately examined their association with T2D risk in the UK Biobank and in an independent, multi-ancestry T2D case:control cohort (AMP-T2D^41^; 20,791 cases, 24,440 controls) to disentangle the effect of GOF variants from the known relationship of LOF variants and increased risk of T2D (Figure 3B). In the meta-analysis, LOF variant carriers compared to neutral variant carriers had an increased risk of T2D as expected from prior studies (OR=1.7, 95% CI=[1.29-2.36], p=2.7 x 10^-4^). Remarkably, GOF variant carriers were protected from T2D (UK Biobank GOF carriers: n=1,464, OR=0.74, 95% CI=[0.59-0.93], p=8.5 x 10^-3^), a signal that was strengthened when analyzed in combination with the AMP-T2D cohort (OR=0.7, 95% CI=[0.62-0.91], p=3.6 x 10^-3^; Figure 3B) evidentiating that genetically determined increases in HNF1A function have clinical consequences in humans.

**Figure 3.**
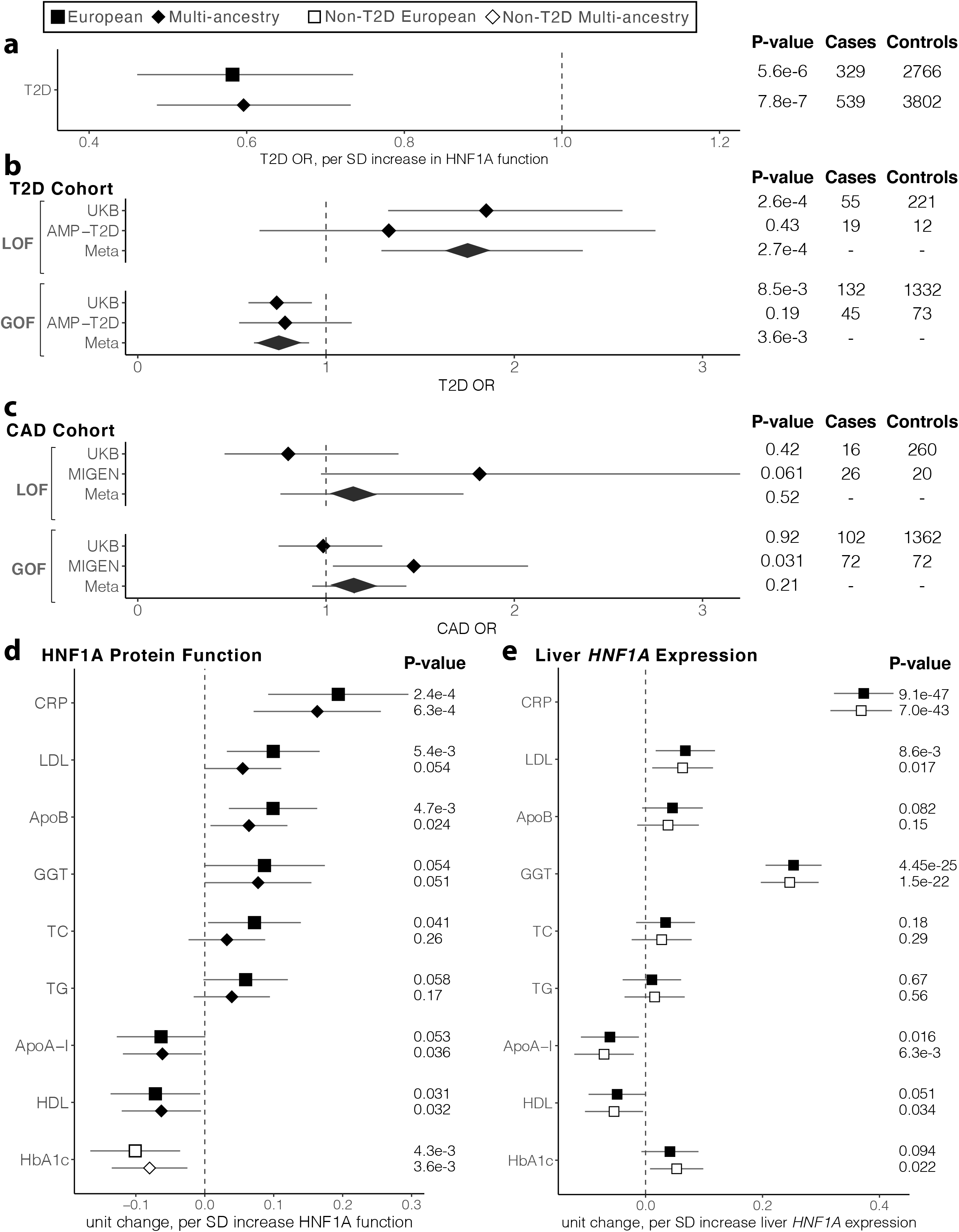
HNF1A GOF variant carriers in the population are protected from T2D, but not CAD, and show elevated levels of serum inflammatory/atherogenic risk factors. Shape and fill indicate the ancestry and T2D status respectively of individuals included in each analysis. All phenotypes are adjusted for age, age^2^, sex, and the first 10 principal components of ancestry. T2D, type 2 diabetes; SD, standard deviation; CAD, coronary artery disease; LOF, loss-of-function; GOF, gain-of-function; CRP, C-reactive protein; LDL, low-density lipoprotein cholesterol; ApoB, apolipoprotein B; GGT, gamma-glutamyl transferase; TC, total cholesterol; TG, triglycerides; ApoA-I, apolipoprotein A-I; HDL, high-density lipoprotein cholesterol; HbA1c, glycated hemoglobin. **a**) Association of HNF1A function score and T2D risk using logistic regression in the UK Biobank *HNF1A* rare protein-coding variant carriers identified among the 454,756 exome sequenced individuals. Odds ratios and the 95% confidence interval are shown. **b**) Association of LOF and GOF HNF1A variants with T2D in the UK Biobank and AMP-T2D (n=20,791 cases, 24,440 controls) using logistic regression. Odds ratios and the 95% confidence interval are shown, and fixed effects inverse-variance meta-analysis was used to combine the results. **c**) Association of LOF and GOF HNF1A variants with CAD in the UK Biobank and the Myocardial Infarction Genetics (MIGen) Consortium (n=24,337 cases, 28,922 controls) using logistic regression. Fixed effects inverse-variance meta-analysis was used to combine the results. **d**) Association of functional HNF1A variants with cardiovascular disease risk factors in the UK Biobank rare variant carriers (European n=3,089, Multi-ancestry n=4,302, linear mixed effects regression). Effect size and the 95% confidence interval are shown for each phenotype. e) Association of liver specific predicted *HNF1A* gene expression with disease risk factors tested in d) among all UK Biobank European genotyped individuals (n=407,227, filled squares) and non-T2D European genotyped individuals (n=376,043, open squares).

Next, we evaluated the association between HNF1A function and CAD events. Since the UK Biobank was underpowered to perform CAD risk analysis due to low incidence of myocardial infarction in this population-based cohort (118 myocardial infarction cases in *HNF1A* rare variant carriers; LOF=16, GOF=102), we jointly analyzed a cohort of individuals exome sequenced for early-onset myocardial infarction (MIGen^42^; 24,337 cases, 28,922 controls). We observed no change in CAD risk in LOF *HNF1A* variant carriers (OR=1.15, 95% CI=[0.76-1.73], p=0.52) nor any decrease in CAD risk in GOF *HNF1A* variant carriers (OR=1.15, 95% CI=[0.93-1.43], p=0.21) even though GOF variant carriers are protected from T2D, an independent CAD risk factor^43^ (Figure 3C). Our combined CAD cohort analysis had over 90% power to detect a decrease in risk of similar magnitude (∼25%) as observed with T2D and GOF variant carriers at the nominal type 1 error rate (α = 0.05).

To explore physiological mechanisms underlying the potentially discordant effects of GOF *HNF1A* variants on T2D and CAD, we quantitatively examined clinical CAD risk factors including hsCRP^16^, lipoprotein levels^44^, liver enzymes^45^, and glycemic traits^46^ in *HNF1A* rare variant carriers in the UK Biobank by regressing the normalized values for each clinical risk factor against the HNF1A function scores of the variant carriers (Figure 3D). A positive relationship was identified between HNF1A function and serum hsCRP, gamma glutamyltransferase (GGT), serum low-density lipoprotein (LDL) cholesterol, and apolipoprotein B (ApoB), while an inverse relationship was identified between HNF1A function and serum high-density lipoprotein (HDL) cholesterol and apolipoprotein A (ApoA). After excluding carriers with diagnosed diabetes, we examined the relationship between HNF1A function and glycemia, finding an inverse relationship between HNF1A function and glycosylated hemoglobin (HbA1c; Figure 3D). These findings suggest that genetic variants which increase HNF1A protein function increase systemic inflammation (hsCRP), oxidative stress (GGT), and promote an atherogenic lipoprotein profile by raising serum ApoB/LDL cholesterol levels and lowering ApoA/HDL cholesterol. However, in concordance with conferring protection from T2D, HNF1A function increasing variants also decrease blood glucose in non-diabetic individuals.

To evaluate if these findings were the result of perturbed HNF1A activity in the liver we turned to liver-specific HNF1A gene expression reasoning that increasing the levels of intra-cellular HNF1A would produce a similar clinical effect as increasing intrinsic HNF1A protein function. To quantify the heritable component of HNF1A gene expression we selected common, non-coding SNPs identified in European liver samples from the Genotype-Tissue Expression (GTEx) Project^47,48^ as expression quantitative trait loci (eQTLs) for HNF1A and combined them to produce a liver HNF1A gene expression score. After removing the carriers of rare protein-coding *HNF1A* variants shown in Figures 3A-D, we assigned liver-specific *HNF1A* expression scores to the remaining 407,227 European individuals using established methods (Functional Summary-based Imputation: FUSION)^49^. These liver-specific *HNF1A* expression scores were then regressed against the same clinical risk factors as the rare, protein-coding variant analysis (Figure 3D). This method confirmed that increased HNF1A gene expression encoded by liver eQTLs conferred increased levels of hsCRP (p < 10^-46^; Figure 3E) as would be expected given the known biology of CRP being synthesized and secreted by hepatocytes^50^. Similarly, serum GGT and LDL cholesterol were associated with increased liver-specific *HNF1A* expression scores, whereas HDL and ApoA showed a nominal inverse association. Notably, glycemia measured by HbA1c (p = 0.09) did not associate with liver-specific HNF1A expression as would be expected from the known pancreatic role of HNF1A in regulating blood glucose^51^.

To evaluate if these associations were being driven by or distinct of T2D, we repeated the above analyses excluding all 41,923 individuals (including 539 carriers of rare-protein coding variants) with diabetes from the UK Biobank population and recomputed the regression (Figure 3D and 3E, Supplementary Table 4) of clinical risk factors against HNF1A function/expression. With regard to liver-specific HNF1A gene expression (n = 376,043 samples without T2D) the analyses were well powered and showed similar effect size, directionality, and significance of all risk factor associations suggesting a causal relationship with HNF1A that is independent of T2D (Figure 3E). Effect sizes in the rare, protein-coding variant analysis also remained directionally consistent, but several associations no longer passed the nominal threshold of statistical significance likely due to a loss of power (n = 3,802 rare-protein coding variant carriers without T2D; Supplementary Table 4). In summary, our data indicate that increased hepatic function/expression of HNF1A increases serum hsCRP, GGT, LDL while decreasing HDL/ApoA independently of its effects on T2D, and increased HNF1A function decreases blood glucose, but not through the liver.

### HNF1A transcriptionally regulates pro-atherogenic gene expression programs in the liver including lipoprotein metabolism, coagulation, and innate immunity genes

To understand the molecular mechanisms by which functional genetic variants in *HNF1A* regulate serum lipids, we deleted *HNF1A* in human hepatocytes using CRISPR/Cas9 and transcriptionally profiled the resulting cells using mRNA sequencing (Figure 4A, Supplementary Table 5). Experiments were carried out in both HUH7 and Hep3B cell lines to mitigate confounding cell line artifacts. Many of the top differentially expressed genes are known transcriptional targets of HNF1A including *FABP1*^39^, *FGB*^52^, and *TM4SF4*^22^. To examine which molecular pathways were perturbed by the loss of *HNF1A*, we performed gene set enrichment analysis (GSEA)^53,54^ on the full list of differentially expressed genes in WT and *HNF1A* KO cells (Figure 4B, Supplementary Figure 6, Supplementary Table 6). The GSEA analysis showed that *HNF1A*-null cells downregulated multiple lipid metabolism pathways including “fatty acid metabolism” (MsigDB #M5935), “cholesterol homeostasis” (MsigDB #M5892), and “bile acid metabolism” (MsigDB #5948), consistent with previous findings from murine *Hnf1a* LOF models^55^. Additionally, pathways for “coagulation” (MsigDB #M5946) and “complement” (MsigDB #M5921) were also significantly downregulated in the absence of HNF1A and a more refined examination of sub-pathways underlying these further showed a downregulation in “platelet activation” (MsigDB #M10857) and “wound healing” (MsigDB #M12074).

**Figure 4.**
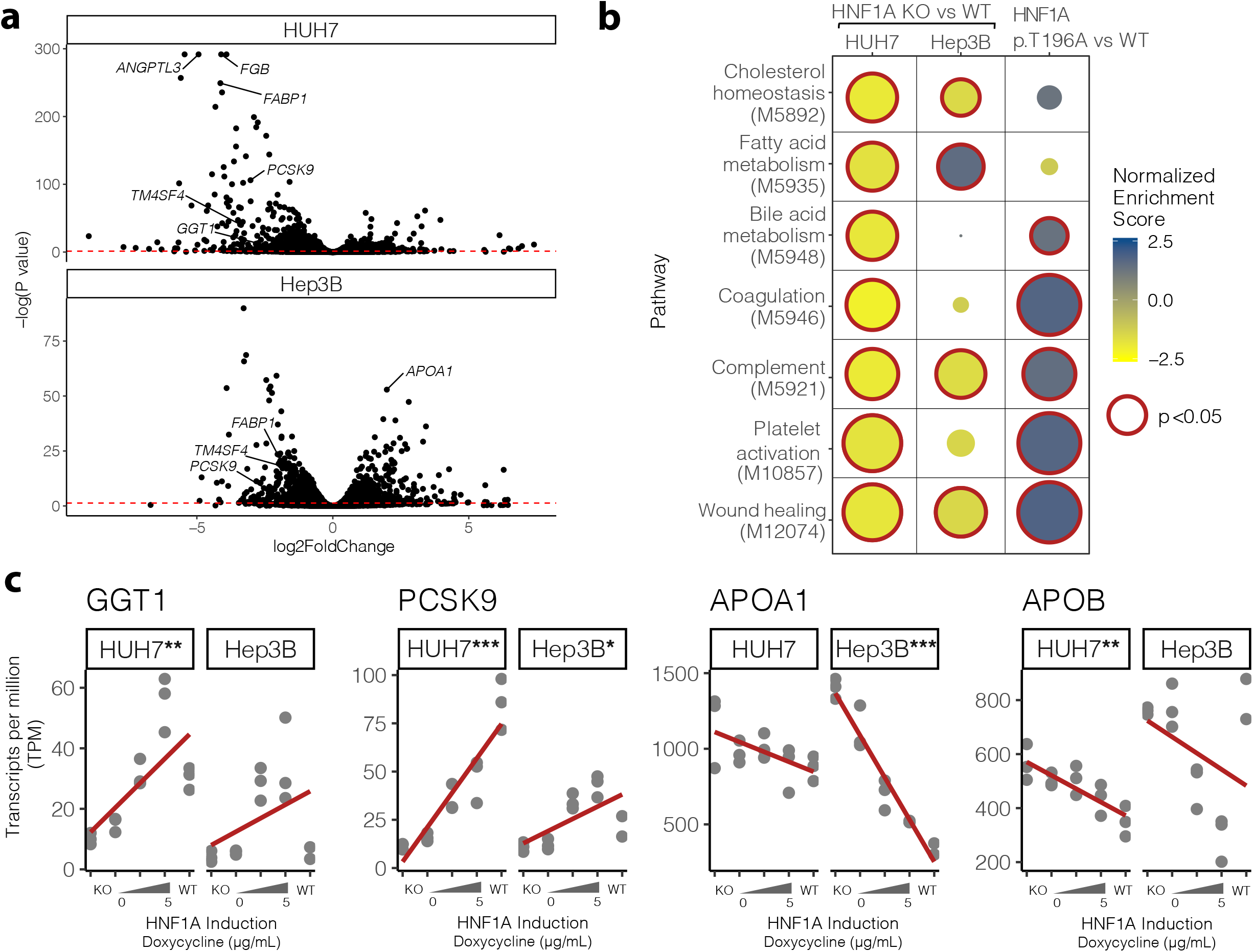
HNF1A transcriptionally regulates lipid metabolism, coagulation, and innate immunity genes. **a)** Differentially expressed genes between wild-type and *HNF1A* knockout (KO) HUH7 and Hep3B cell lines quantified by mRNA sequencing (n=3 per group). The dotted red line indicates an adjusted p-value threshold of 0.05, with genes falling above the line meeting a global significance threshold for differential expression. **b**) Gene set enrichment analysis (GSEA) was performed on mRNA sequencing data shown in **a)** to identify pathways altered by deletion of HNF1A (HNF1A KO vs WT). Significant genesets in at least one cell line are shown. The effect of gain-of-function (GOF) in HNF1A on these pathways was examined by complementing HNF1A KO HUH7 cells with WT HNF1A and the GOF p.T196A variant (n=3 per group) followed by mRNA sequencing, differential expression, and GSEA (HNF1A p.T196A vs WT). Downregulated pathways (negative normalized enrichment scores) are shaded yellow and upregulated pathways are shaded blue. Size represents the -log10(P-value), and significantly enriched pathways (p<0.05) are outlined in red. **c**) *HNF1A* was reintroduced via doxycycline inducible transgenes into *HNF1A* KO hepatocyte cell lines and global gene expression was measured (TPM = transcripts per million, n=3 per group). In a dose dependent fashion, HNF1A regulates genes involved in oxidative stress and lipid regulation pathways. Significance levels from regressing TPM on HNF1A levels are shown for each gene and cell type if significant (*** p <10^-6^, ** p<10^-3^, * p<0.05, linear model). (*GGT1*, Gamma Glutamyltransferase 1; *PCSK9*, Proprotein Convertase Subtilisin/Kexin Type 9; *APOA1*, Apolipoprotein A1; *APOB*, Apolipoprotein B)

To evaluate global transcriptional changes from *HNF1A* GOF variants, we assessed the global transcriptional effects of expressing HNF1A WT and the most commonly found human GOF variant HNF1A p.T196A (Figure 2B). We performed short-term complementation experiments, expressing HNF1A WT and p.T196A in *HNF1A*-null HUH7 cells for 18 hours from in vitro-transcribed mRNA at levels similar to endogenous HNF1A in WT HUH7 cells (Supplementary Figure 7). GSEA analysis of genes differentially expressed between the HNF1A WT- and p.T196A variant-complemented cells (Supplementary Table 7) showed that relative to WT, the p.T196A variant more strongly upregulated several of the lipid metabolism and coagulation/complement activation pathways that were downregulated in *HNF1A* KO cells (Figure 4B, Supplementary Table 6), supporting a molecular mechanism of GOF that spans multiple transcriptional pathways.

To validate specific effector genes that could explain the observed inflammatory, oxidative stress, and atherogenic serum lipoprotein gene expression profiles in *HNF1A* variant carriers, we performed independent experiments evaluating HNF1A:effector gene dose responsiveness. Doxycycline-inducible *HNF1A* transgenes were introduced into *HNF1A*-null cells and exposed to varying doses of doxycycline to evaluate HNF1A:gene dose-response curves, quantified by regressing gene transcripts per million (TPM) onto HNF1A dose (Figure 4C). The most straightforward example was for serum GGT, a marker of oxidative stress^56^ which is hepatically secreted, positively regulated by HNF1A (*GGT1* p < 10^-3^; HUH7 Figure 4C), and whose serum values are positively correlated with genetically mediated increases in HNF1A function/liver-specific gene expression (Figure 3D and 3E). Effector genes that could account for the serum lipoprotein profile associated with GOF in HNF1A and showed dose response included the LDL receptor recycling regulator Proprotein Convertase Subtilisin/Kexin Type 9^57^ (*PCSK9* p < 10^-6^, HUH7) and apolipoprotein A1, the major lipoprotein component of HDL (*APOA1* p < 10^-6^, Hep3B). Conversely, a negative HNF1A:gene expression dose response was observed for apolipoprotein B, the major lipoprotein component of LDL (*APOB* p < 10^-3^; HUH7) suggesting the positive correlation observed between serum ApoB levels with genetically mediated increases in HNF1A function/expression (Figure 3D and 3E) is not a hepatocyte cell autonomous effect of HNF1A.

To corroborate direct transcriptional regulation of these genes by HNF1A, we identified HNF1A consensus binding motifs within a 5 kilobase genomic window of the transcription start sites (TSS) and overlaid these with HNF1A ChIP-seq data from HepG2 cells made available through the ENCODE project^58^. The *PCSK9 and GGT1* loci showed ChIP-seq peaks coincident with high-scoring HNF1A consensus motifs near the TSS (Supplementary Table 8) strongly supporting direct transcriptional regulation. The *APOA1* and *APOB* loci showed more modest evidence of direct regulation having only ChIP-seq peaks near the TSS but lacking HNF1A consensus motifs.

## Discussion

In this study we combine massively parallel functional characterization and saturation mutagenesis with genetic and clinical data from almost 600,000 individuals to evaluate a full allelic series of protein-coding variants in *HNF1A* and their impact on human health. By correlating variant, cellular function, and clinical phenotype in thousands of individuals, we find that the general human population contains carriers of gain-of-function (GOF) variants that increase the transcriptional activity of HNF1A by 30-50%. These GOF variants have both beneficial and pathological clinical consequences in the people who carry them through pleiotropic actions in the pancreas and liver respectively. While GOF variants confer a ∼30% decrease in T2D risk they, likely through liver-specific transcriptional effects, increase levels of serum CAD risk factors and cause an atherogenic lipid profile (increased serum ApoB/LDL and decreased ApoA/HDL). Lastly, we demonstrate that the hepatic transcriptional regulation of key effector genes by HNF1A, including *APOA1* and *PCSK9*, mediates at least part of the proatherogenic lipid profile observed in GOF variant carriers.

To our knowledge this study provides the first description of function increasing genetic variants in *HNF1A* and their presence in the human population, an unexpected finding given that thousands of individuals have undergone *HNF1A* gene sequencing and dozens of protein-coding variants have been functionally characterized over the past two decades^59,60^. We note that in contrast to LOF variants, GOF HNF1A variants showed no evidence of being under evolutionary constraint (Supplementary Figure 3) and thus would escape discovery by computational prediction programs^27,28^ which rely on evolutionary conservation, highlighting the importance of taking an experimental approach to pathogenicity prediction at clinically important genes. In retrospect, some variants we have demonstrated as GOF have been previously identified and experimentally tested showing transactivation signals greater than WT^37^, but due to inconsistent results across different experiments or labs were labeled WT-like. For example, the p.T196A variant showed activity greater than WT in transformed COS-7 monkey fibroblasts but not in mouse insulinoma cells^37^ whereas in a recent study^61^ p.P291Q and p.T196A showed increased transcriptional activity over WT in both HeLa and rat insulinoma cells but at only one of two expert labs testing the same variants. In our own experiments we were able to most clearly resolve variants with function greater than WT in cell systems without endogenous HNF1A (HUH7 KO and Hela cells; Figure 2C), but the signal was more challenging to identify in MIN6 cells with a high level of basal HNF1A activity (Figure 2D). Moreover, such GOF variants, by virtue of conferring protection from diabetes, were less likely to be identified in previous studies that selected diabetic individuals for sequencing. By functionally characterizing all possible missense variants in a single experiment, our study revealed the full spectrum of possible functional variation including both loss and gain. We further benefited from ascertainment of *HNF1A* variants in the UK Biobank, a general population-based cohort not enriched for diabetes and therefore not depleted of GOF *HNF1A* variant carriers. In the context of another recent study that identified GOF variant carriers in MC4R in the UK Biobank^62^, our study highlights the potential for discovering novel genetic mechanisms of disease by conducting large scale functional variant characterization and clinical correlation in population-based cohorts.

Through the identification of a full allelic series of HNF1A variants, our study advances previously reported genetic associations between HNF1A, clinical risk factors and metabolic diseases (T2D^40^ and CAD^14^) by providing new tissue-specific, mechanistic insights and unexpected directional relationships. In the general population our data support that GOF in HNF1A reduces T2D risk and glycemia through enhanced beta cell function which complements the mechanisms by which LOF HNF1A variants are known to cause T2D/MODY3^3,35^. In the liver, however, we find that GOF in HNF1A is metabolically pathogenic causing increases in hepatically derived lipoprotein risk factors which have been shown causal for CAD as well as increases in correlative if not directly causal inflammatory (hsCRP), and oxidative (GGT) risk factors^16,45, 63–65^. Despite a significant decrease in diabetes risk in GOF variant carriers, we found no signal for protection from CAD risk in GOF variant carriers and propose that the increase in hepatic risk factors is counterbalanced by the decrease in T2D risk (Figure 5). While glycemic and inflammatory factors may influence CAD risk through insulin resistance and diabetes^66,67^, our analyses support that GOF in HNF1A acts independently of diabetes (Figure 3D and 3E). An important medical implication of these findings is that HNF1A ‘replacement’ gene therapy for HNF1A MODY patients, which is technically feasible through mRNA therapeutics^68^, should be approached with caution as hepatic increases in HNF1A would be clinically undesirable.

**Figure 5.**
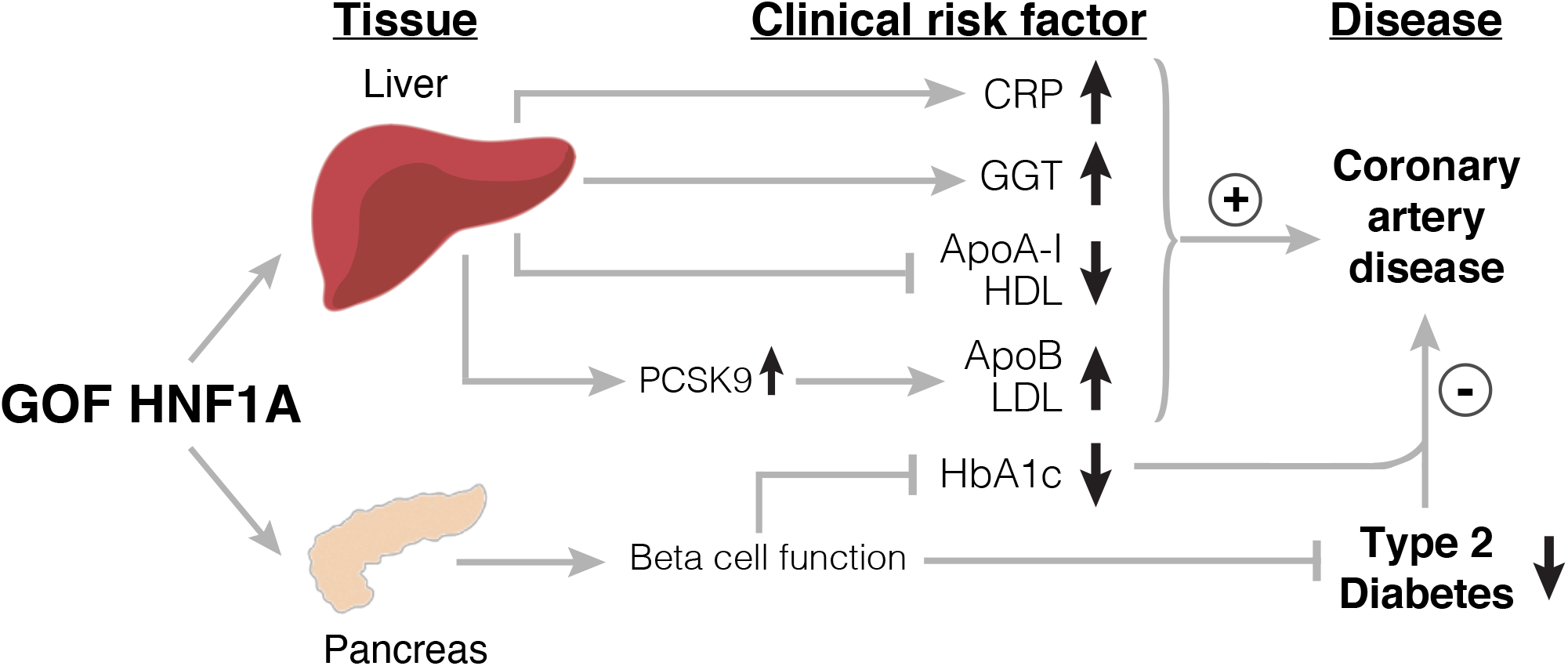
Graphical summary of proposed mechanism through which gain-of-function in HNF1A decreases diabetes risk through pancreas-specific mechanisms but increases the hepatic secretion of other independent CAD risk factors. CAD, coronary artery disease; T2D, type 2 diabetes; GOF, gain-of-function; CRP, C-reactive protein; GGT, gamma-glutamyl transferase; ApoA-I, apolipoprotein A-I; HDL, high-density lipoprotein cholesterol; PCSK9, proprotein convertase subtilisin/kexin type 9; ApoB, apolipoprotein B; LDL, low-density lipoprotein cholesterol; HbA1c, glycated hemoglobin. Carriers of GOF variants in HNF1A have reduced T2D risk and HbA1c through enhanced beta cell function but have increases in multiple other independent CAD risk factors including serum inflammation (CRP), oxidative stress (GGT), and proatherogenic lipids (elevated LDL through increased transcription of *PCSK9* and decreased HDL through decreased transcription of *APOA1*). GOF in HNF1A increases hepatic secretion of CAD risk factors, but CAD event risk is attenuated by a counterbalancing decrease in T2D risk.

Molecularly, we find that the alterations in serum inflammation (hsCRP) are concordant with pro-inflammatory gene expression changes in complement and coagulation factors secreted by the liver (Figure 4) which may be the underlying causal CAD risk mediators. Regarding serum lipid alterations, murine LOF studies have implicated HNF1A in bile acid secretion and cholesterol metabolism^55^, findings corroborated in humans by our serum biomarker and cellular data (Figures 3 and 4). At the gene level, our data suggest that the suppression of ApoA/HDL levels by HNF1A is likely cell-autonomous as it occurred in cultured hepatocytes. We propose that the increase in serum LDL cholesterol in carriers of rare HNF1A GOF variants is at least partially mediated by a direct transcriptional activation by HNF1A of PCSK9 (Figures 4 and 5), a hepatic protease which regulates hepatic uptake of LDL^69,70^ and is a pharmacological target for lipid lowering therapies^71–73^. The lack of an HNF1A motif at the *APOB* promoter as well as the lack of positive HNF1A-APOB dose:response (Figure 4C) in cultured hepatocytes suggests the increase in serum ApoB levels from genetically mediated increase in HNF1A function/expression is indirect possibly through increased serum LDL persistence that would be caused by increasing PCSK9 transcription^74^ (Figure 5). Intriguingly, our study predicts that GOF variant carriers in *HNF1A*, by virtue of enhanced PCSK9 activity, may be a subpopulation that may respond to PCSK9 inhibitors more favorably for LDL reduction than conventional statin medications which are known to increase levels of PCSK9^75^.

Limitations of our study include the precision of HNF1A function scores generated in our massively parallel assay and the cell models utilized to elucidate molecular mechanisms. While our observation of a new class of amino acid substitutions that increase the transcriptional activity of HNF1A (Figure 1D and 2A) is strongly supported by experimental validation in multiple independent experiments (Figure 2C, 2D, 2E, and 4B), the precise level of increase in function conferred by any particular variants contains biological (different cellular contexts) and experimental (performed in different labs) uncertainty that limit their interpretation. For example p.P291Q was the top scoring GOF variant (Figure 2A), replicated in HeLa cells (Figure 2C) but did not show increase over WT in MIN6 cells (Figure 2D) or ANGPTL3 secretion (Figure 2E) assays. Conversely, p.T196A was the lowest scoring of the selected GOF variants but showed higher function than other variants in subsequent assays. Thus, we would caution against utilizing these function scores directly to provide an individualized risk estimate to a person carrying a protein-coding *HNF1A* variant. If used as a standalone classifier for GOF/LOF, our function scores would meet the 90% probability standard for clinical variant interpretation^76^ for only 43 variants (Supplementary Table 2). Additionally, due to the design of our deep mutational scan, effects of splice variants are not detectable and functional interpretation of such variants should be approached with caution. Furthermore, our mechanistic evaluation in multiple human hepatocyte cell lines could miss important regulatory mechanisms due to the limitations of *in vitro* cell culture models. For example, our cellular data did not identify direct regulation of CRP by HNF1A as it is poorly expressed and lacks evidence for binding in the publicly available HNF1A ChIP-seq data^58^ (Supplementary Table 8), but previous investigators have reported HNF1A binding at the CRP promoter^77–79^ supporting a direct regulation.

In summary, we find that ∼1:300 individuals in the general population carry GOF variants in *HNF1A* which are not identifiable by computational prediction tools and pleiotropically decrease diabetes risk while increasing hepatic secretion of CAD risk factors. In an era of rapidly deployable gene therapy^80^, understanding the clinical consequences of increasing or decreasing gene function in humans bottlenecks therapeutic development. As biobanks continue to grow in size and phenotypic complexity, this study demonstrates the utility of comprehensive functional characterization to distill novel mechanisms of disease and identify adverse effects prior to undertaking drug development and putting clinical trial participants at risk.

## Author Contributions

N.D and A.R.M. designed the study. N.D., B.K., S.Hu, R.I., M.W., J.G., X.D., C.D.A.S., and A.M.D. generated/acquired and analyzed the data. N.D., B.K., S.Hu, R.I., M.W., R.A., J.F., G.M.P., S.H., P.G., A.M.D., A.V.K., J.O., V.R., and A.R.M., were involved in data interpretation. N.D. and A.R.M. drafted the manuscript. N.D., B.K., R.I., M.W., X.D., V.M., G.M.P., P.G., A.M.D., A.V.K., J.O., V.R., S.H., and A.R.M. were involved in critical manuscript revision.

## Data / Code Availability

All UK Biobank data used in this study are accessible through application to UK Biobank. Data and code to be made available upon manuscript acceptance.

## Methods

### Cell line generation

HUH7 and Hep3B cells were cultured in DMEM (Life Technologies #11995065) supplemented with 10% FBS (Sigma #F2442) and L-Glutamine and antibiotics (Omega PG-30). Polyclonal *HNF1A*-null cells were generated by introducing the endonuclease Cas9 and sgRNA (CTGTCCCAACACCTCAACAA) targeting exon 2 of human *HNF1A* into cells by lentiviral transduction as we have previously described^19^.

### Synthesis of all possible HNF1A amino acid variants

Human *HNF1A* cDNA (CCDS9209.1) was recoded by selecting alternative codons to eliminate the CRISPR/CAS9 binding site and optimize expression (Supplementary Table 9). This cDNA corresponds to the longest amino acid coding sequence of HNF1A expressed in the liver and also the most abundant^48^. Constructs encoding all 19 alternative amino acids at each position were synthesized (Twist Bioscience, Supplementary Table 1). An additional 613 constructs encoding synonymous variants spread across the coding sequence were also synthesized and included. The HNF1A construct library described above was cloned into a lenti-viral expression vector by simple restriction cloning, sequence verified for quality and completeness (Supplementary Figure 1), and transfected into a packaging cell line to produce pooled lenti-virus^19^.

### Pooled experimental assay of 11,970 HNF1A variants

The lentiviral library of HNF1A amino acid variants was introduced into HUH7 *HNF1A*-null cells under a doxycycline responsive promoter to enable controlled protein expression^81^. To minimize doubly infected cells, 1.2x10^7^ viral particles were combined with 4x10^7^ cells after which uninfected cells were eliminated by selection with 4 μg/mL puromycin. At least 10^7^ cells were infected to ensure that each HNF1A variant was independently represented in 1000 cells. The resulting polyclonal population of HUH7 hepatocytes (each cell containing a different HNF1A variant) was stimulated with doxycycline (5 µg/mL) for 6 days and then immuno-stained for TM4SF4 (R&D systems FAB7998A). Stained cells were sorted using FACS (FACS Aria II and BD Influx) into two equal bins: high expression and low expression bins based on TM4SF4 expression with a ten-fold expression intensity difference between each bin (Figure 1A). Control experiments with *HNF1A*-null and WT cells were performed to identify the TM4SF4 expression distributions of cells with and without HNF1A (Figure 1A dashed red and dashed green lines and Supplementary Figure 2). These distributions helped to establish TM4SF4 expression cutoffs for the FACS sorting. Three independent sorting experiments were performed, sorting ∼500,000 cells per bin per replicate. To re-identify and quantitate the HNF1A variants in the TM4SF4 ‘high’ and ‘low’ bins, genomic DNA (gDNA) was extracted (Qiagen #51304) from each sample of 500,000 cells and the integrated proviral HNF1A transgenes recovered by PCR. Each sample of purified gDNA was split into 14 separate 100 uL PCR reactions (∼400ng gDNA per reaction) and amplified using Herculase II (Agilent #600675) according to the manufacturer protocol using a Tm of 57°C (see Supplementary Table 9 for primer sequences). All 14 PCR reactions per sample were pooled together, and the transgenes were purified (Qiagen #28104) and prepared for sequencing (Illumina Nextera #FC-131-1096) according to the manufacturer’s protocol. The six samples were pooled and sequenced to obtain approximately 350 million paired-end 100-base reads (Illumina NovaSeq) resulting in a 12x base coverage assuming a target size of 6 x 10^9^ bp (2000 bp transgene x 12,000 variants x 250 observations per variant). Raw sequencing reads were aligned to the reference HNF1A recoded cDNA sequence (Supplementary Table 9) using ORFCall (https://github.com/tedsharpe/ORFCall)^19^ and the number of occurrences of each amino acid at each position along the coding region were counted and tabulated.

### Calculation of function scores

We constructed a likelihood function based on the log odds of an amino acid variant being in the fraction with high or low TM4SF4. The log odds for each amino acid variant was estimated by maximizing a likelihood function based on the observed counts of each amino acid variant in the fractions with high and low TM4SF4 normalized to the total read depth at that amino acid position. Data were combined across experimental replicates. To avoid spuriously high or low log-odds estimates for any given variant, we constrained the log-odds estimate with a Gaussian prior whose parameters were estimated from data combined across all variants. These procedures were analogous to those we have previously described^19^. Variants which had total counts across all replicates and bins less than the known low count variants from the original library design were not assigned a function score (125 variants).

To calculate the posterior error probability (PEP) that any particular variant was not neutral, we computed the probability that it belonged to the synonymous function score distribution instead of the nonsynonymous distribution using the formula PEP_nonWT = P_synonymous_/(P_synonymous_ + P_nonsynonymous_)^29^. The probability density functions for P_synonymous_ and P_nonsynonymous_ were generated by linearly interpolating the distribution of function scores (“combined_function_score” Supplementary Table 2) for synonymous and nonsynonymous variants using an interpolation function as instantiated in R v3.5.1.

### Genetic data

UK Biobank research was conducted under application numbers 51436 and 26041. For all exome sequenced cohorts, variants within the genomic coordinates of *HNF1A* (chr12:120,977,683-121,002,512, GRCh38) were extracted, and variant annotation was performed using SnpEff v4.3^82^. Nomenclature used for missense variants is for the canonical HNF1A transcript ENST00000257555.10; protein ENSP00000257555.4. Disruptive variants (i.e. splice region variants, stop gained variants, or frameshift variants) were assigned the function score equal to the lowest known pathogenic variant functional score, homozygous carriers of rare HNF1A protein-coding variants (n=5) were removed from all analyses, and compound heterozygotes (n=39) were included twice as single variant carriers.

### Metabolic biomarkers and CAD risk factors

Clinical measurements from all 502,489 UK Biobank participants were log transformed, if not normally distributed (Supplementary Table 4) and residualized adjusting for age, age^2^ and sex, then normalized using a rank based inverse normal transformation^83^. Diabetes mellitus designation was based on a compilation of self-report, use of insulin, ICD-10 codes of E10.X, E11.X, E12.X, E 13.X, E14.X in hospitalization or primary care records^84^, or HbA1c levels greater than or equal to 48 mmol/mol (6.5%)^85^.

### Genotype-function-phenotype association analyses

To examine the association between the full continuum of HNF1A function score and diabetes status in the UK Biobank, we performed logistic mixed effects regression (lme4 v1.1-17, lmerTest v3.1-2) with variant identity as a random effect in order to account for multiple carriers of the same variant, adjusting for age, age^2^, sex, and the first 10 genetic principal components of ancestry (computed by UK Biobank for multi-ancestry analyses (Field 22009) and computed for European ancestry participants as previously described^86^). To examine the association between function bins (i.e. GOF/LOF) and diabetes or CAD status in the UK Biobank, we performed logistic regression (glm function, R stats v3.5.1) adjusting for covariates previously mentioned. To account for related individuals, in the full multi-ancestry analysis we randomly removed one participant if a pair of them had a genetic relationship equal or closer than a second-degree relative, and in the European analysis, related individuals were identified and excluded as per Deaton et al^86^.

Analysis in the AMP-T2D-GENES study was conducted as previously described^41^. Briefly, we conducted two burden tests -- one including gain of function variants and one including loss of function variants -- using the EPACTS software package, regressing sample phenotype on genotype dosage. We conducted each test across all unrelated individuals pooled together and included principal components of ancestry covariates (computed from common variants) as well as covariates for sample cohort of origin and sequencing technology. The effect size and significance of the burden test was evaluated using the two-sided Firth logistic regression test. Meta-analysis for the UK Biobank and AMP-T2D cohorts was conducted using fixed-effect inverse-variance meta-analysis as implemented by METAL^87^.

To test the association of rare *HNF1A* variants in the MIGen ExSeq data set with the risk of coronary artery disease, a Firth logistic regression was applied with sex and top 10 principal components of ancestry as covariates and cleaned to second-degree of relationship. The Firth logistic regression is a test robust to association testing in the context of low carrier counts or case-control imbalance^88^. For each person, the genetic score of the qualified rare variant carriers was coded as 1, and 0 otherwise. Meta-analysis for the UK Biobank and MIGen ExSeq cohorts was conducted using fixed-effect inverse-variance meta-analysis as implemented by METAL^87^. Power calculations were performed using the Purcell et. al.^89^ genetic power calculator with the following parameters: high risk / marker allele frequency = cumulative GOF allele frequency = 0.002, a 5.4% prevalence of CAD in the UK Biobank, genotype relative risk = GOF OR for T2D = 0.75, total number of CAD cases in MIGEN and UK Biobank = 49,666, D’ = 1, and control:cases ratio = 9.44. Power is reported for α = 0.05.

Cardiovascular disease risk factor phenotypic associations with function scores were performed within the UK Biobank using the normalized clinical measurements described above and a linear mixed model with function score as a fixed effect, adjusting for 10 genetic principal components of ancestry as described above, and with variant identity as a random effect in order to account for multiple carriers of the same variant (lme4 v1.1-17, lmerTest v3.1-2).

To quantify liver specific HNF1A gene expression, the Functional Summary-based Imputation (FUSION) framework for generating individual-level predicted gene expression^49^ was applied to UK Biobank genotyped individuals. First, to ensure that this analysis was independent of our analysis of protein-coding variants, carriers of rare protein-coding HNF1A variants (n=2,203) in the UK Biobank were excluded. Next, all HNF1A (ENSG00000135100) expression reference weights computed in European liver RNA-sequenced samples from the Genotype-Tissue Expression (GTEx) Project v8 were extracted from the FUSION archive (http://gusevlab.org/projects/fusion/#gtex-v8-multi-tissue-expression) (n=466 SNPs with precomputed expression weights, Supplementary Table 10). These expression weights and their corresponding imputed genotypes in the UK Biobank European individuals (n=407,227) were combined into a predicted gene expression score for each individual by computing a linear combination across the HNF1A SNPs expression weights and genotypes, then dividing by the number of non-missing SNPs as implemented in the Plink score utility^90^. Phenotypes were normalized as described above and regressed onto predicted HNF1A liver expression scores using a generalized linear model (glm function, R stats v3.5.1) adjusting for 10 genetic principal components of ancestry. Regressions additionally included as a covariate individual level genotype of the common HNF1A variant rs1169288 (encoding p.I27L) which has previously been associated with metabolic phenotypes^14,91^ and was significantly associated with the predicted HNF1A liver expression score (p < 2 x 10^-16^, linear model).

### Functional validation of GOF HNF1A variants

Each GOF variant selected for validation (Figure 2A) was recreated as a transgene based on the native sequence of the cDNA (CCDS9209.1) with the exact human carrier nucleotide change observed in the UK Biobank (Supplementary Table 9). The constructed variants were tested for transactivation potential in HeLa cells as we have previously published^36^ using the rat albumin promoter. All transactivation experiments included three parallels performed on three independent experimental days (nine readings in total).

Transcriptional activity of the HNF1A GOF variants was also tested in MIN6 cells using the HNF4aP2 promoter^37^. Cells were co-transfected with a DNA mixture consisting of 10 ng of pGL-Luc and pGL4.75, and 10 ng of variant or WT *HNF1A*. The level of the transcription factor activity was evaluated by measuring firefly luciferase activity relative to renilla luciferase activity using the Dual Luciferase Assay System (Promega #E1500), as described by the manufacturer.

### Assessment of ANGPTL3 levels in cultured hepatocytes

HNF1A constructs (WT, p.T260M, and p.P447L) were introduced into WT and *HNF1A* KO human HUH7 cells by lentiviral induction under a doxycycline responsive promoter as above. Cells were stimulated with doxycycline (0, 0.5, 1, 5 µg/mL) for 5 days. Following doxycycline incubation, the spent media was collected for ANGPTL3 ELISA analysis (R&D Systems #DANL 30) and the cells for protein quantification (RIPA buffer + protease inhibitor) according to the manufacturer’s protocol. GOF variant constructs were transfected into HUH7 hepatocytes using TransIT-LT1 transfection reagent (Mirus Bio), puromycin selection was performed to remove untransfected cells, and cells were incubated in fresh media for 24 hours before harvest for ELISA and protein quantification as described above.

### Functional genomic analysis of HNF1A deleted hepatocytes and complementation with GOF variants

*HNF1A*-null HUH7 and Hep3b cells were generated as described above, and WT HNF1A was reintroduced using a doxycycline-inducible promoter (0, 2, 5 µg/mL) in triplicate. mRNA was extracted (Qiagen #74104) and sequenced (Illumina TruSeq) according to the manufacturers’ protocol to a depth of at least 30 million reads per sample. Raw reads were aligned using Kallisto (v0.44.0)^92^ with default parameters to generate gene counts per cell. DESeq2 (R v3.5.1, package v1.22.1)^93^ was used to perform differential expression analysis with effect sizes and p-values obtained from the Wald test as instantiated within DESeq2. Gene set enrichment analysis (GSEA)^53,54^ was conducted using the fgsea package with 10000 permutations (package v1.8.0^94^) utilizing Hallmark gene sets^95^ and Gene Ontology biological process gene sets^96,97^ related to significantly enriched Hallmark gene sets (cholesterol homeostasis, fatty acid metabolism, bile acid metabolism, coagulation, and complement activation). Significance was determined by a two-sided enrichment p-value with Benjamini–Hochberg correction for multiple hypotheses as instantiated within the fgsea software package.

Complementation of *HNF1A*-null HUH7 cells with mRNA encoding WT HNF1A or the GOF p.T196A variant was essentially performed as described before^98^, with modifications. Briefly, cDNA constructs were amplified from the expression plasmids described above with primers that introduce a T7 promoter and AG as the first two transcribed nucleotides and that add a 179-nt-long poly(A) tail to the 3’ end. PCR products were phenol/chloroform-extracted and in vitro-transcribed with T7 RNA Polymerase HiScribe Mix (NEB E2040S), CleanCap Reagent AG (TriLink N-7113) and 5-methoxyuridine triphosphate instead of UTP for 2 hours at 37°C to generate capped, polyadenylated mRNA. IVT mRNA was precipitated with 2.5 M LiCl final, washed twice with 80% EtOH and dissolved in water. For each construct, 10^6^ *HNF1A*-null HUH7 cells were electroporated with 2 µg mRNA in 100 µl Buffer R with four 10-millisecond pulses at 1230 V using a Neon Transfection System (ThermoFisher MPK10025). Cells were cultured for 18 hours in 5 mL medium in 60 mm plates, and mRNA was extracted (Zymo R1050), sequenced (Illumina Stranded mRNA), and gene expression analyzed as above. Expression of HNF1A was verified by Western blot, and EGFP mRNA was used as electroporation control. Differential expression analysis between *HNF1A*-null HUH7 complemented with EGFP and WT HNF1A showed a profile of differentially expressed genes similar to *HNF1A*-null vs HNF1A WT cells (top genes in both experiments included FGB, FABP1, PCSK9, and ANGPTL3; Supplementary Table 5 and 7)

HNF1A binding motifs were scored using JASPAR^99^ consensus matrix sequence matches 5000 bp before and after the genomic coordinates of the transcriptional start site for each gene of interest. HNF1A HepG2 ChIP-seq bigWig signal p-value and IDR ranked peaks result files from both replicates were downloaded from ENCODE experiment ENCSR633HRJ^100^.

## Data Availability

All UK Biobank data used in this study are accessible through application to UK Biobank.

## Acknowledgements

We would like to thank Joseph Witztum and Vikas Bansal for helpful discussions. We are grateful to Xiaolan Zhang and Kyle Sanchez for assistance with preparing the HNF1A variant library, cellular screening/FACS sorting, and production of *HNF1A*-null hepatocyte cell lines. We would also like to thank Twist Bioscience for constructing the HNF1A variant library. Henrietta Lacks and the HeLa cell line that was established from her tumor cells without her knowledge or consent in 1951 have made significant contributions to scientific progress and advances in human health. We are grateful to Henrietta Lacks, now deceased, and to her surviving family members for their contributions to biomedical research.

## Alnylam Human Genetics

Aimee M. Deaton^1^, Gregory Hinkle^1^, Rachel A. Hoffing^1^, Aaron M. Holleman^1^, Philip LoGerfo^1^, Paul Nioi^1^, Mollie E. Plekan^1^, Lucas D. Ward^1^, Carissa Willis^1^

### Affiliations

^1^ Alnylam Pharmaceuticals, Cambridge, MA, USA

## AMP-T2D

### List of consortium members

Gonçalo R. Abecasis^1,2^, Carlos A. Aguilar-Salinas ^3,4^, David M. Altshuler^5,6,7,8^, Gil Atzmon^9,10,11^, Francisco Barajas-Olmos^12^, Aris Baras^13^, Nir Barzilai^10^, Graeme I. Bell^14^, Thomas W. Blackwell^1^, John Blangero^15^, Michael Boehnke^16^, Eric Boerwinkle^17,18^, Lori L. Bonnycastle^19^, Erwin P. Bottinger^20,21^, Donald W. Bowden^22,23^, Jennifer A. Brody^24^, Brian Burke^25^, Noël P. Burtt^8^, David J. Carey^26,27^, Lizz Caulkins^8^, Federico Centeno-Cruz^12,28^, John C. Chambers^29,30,31,32^, Juliana Chan^33^, Edmund Chan^34^, Ling Chen^35^, Siying Chen^16^, Ching-Yu Cheng^36,37,38,39^, Francis S. Collins^19^, Cecilia Contreras-Cubas^12^, Adolfo Correa^40^, Maria Cortes^41^, Nancy J. Cox^14,42^, Emilio Córdova^12^, Dana Dabelea^43,44^, Paul S. de Vries^17^, Ralph A. DeFronzo^45^, Frederick E. Dewey^13^, Lawrence Dolan^46^, Kimberly L. Drews^25^, Ravindranath Duggirala^15^, Josée Dupuis^47^, Maria Elena Gonzalez^48^, Amanda Elliott^8,35^, Maria Eugenia Garay-Sevilla^49^, Jason Flannick^8,50,51^, Jose C. Florez^5,8,52^, James S. Floyd^53^, Philippe Frossard^54^, Christian Fuchsberger^1,55^, Stacey B. Gabriel^41,56^, Humberto García-Ortiz^12^, Christian Gieger^57,58,59^, Benjamin Glaser^60^, Clicerio Gonzalez^61^, Niels Grarup^62^, Leif Groop^63,64^, Myron Gross^65^, Christopher A. Haiman^66^, Sohee Han^67^, Craig L. Hanis^17^, Torben Hansen^62,68^, Nancy L. Heard-Costa^69,70,71^, Susan R. Heckbert^72^, Brian E. Henderson^66^, Soo Heon Kwak^73^, Anne U. Jackson^74^, Young Jin Kim^67^, Marit E. Jørgensen^75,76,77,78^, Megan Kelsey^25,43^, Bong-Jo Kim^67^, Ryan Koesterer^8^, Heikki A. Koistinen^79,80,81^, Jaspal S. Kooner^31,32,82,83^, Johanna Kuusisto^84,85,86^, Markku Laakso^84,85,86,87^, Leslie A. Lange^88,89,90^, Joseph B. Leader^26,27^, Juyoung Lee^67^, Jong-Young Lee^91,92^, Donna M. Lehman^45^, H. Lester Kirchner^26,27^, Allan Linneberg^93,94,95,96^, Jianjun Liu^34,97,98^, Ching-Ti Liu^47^, Ruth J. F. Loos^20,99^, Valeriya Lyssenko^63,100^, Ronald C. W. Ma^33^, Anubha Mahajan^101^, Alisa K. Manning^8,52,102,103^, Juan Manuel Malacara-Hernandez^49^, Anthony Marcketta^13^, Angélica Martínez-Hernández^12^, Karen Matsuo^16^, Elizabeth Mayer-Davis^104^, James B. Meigs^41,52,103,105^, Thomas Meitinger^106,107,108,109^, Elvia Mendoza-Caamal^12,28^, Josep M. Mercader^8,110,111^, Hyun Min Kang^1^, Karen L. Mohlke^88,112^, Andrew P. Morris^113,114,115^, Andrew D. Morris^116^, Alanna C. Morrison^17^, Anne Ndungu^115^, Maggie C. Y. Ng^117^, Peter Nilsson^118^, Christopher J. O’Donnell^52,53,119,120,121,122,123^, Colm O’Dushlaine^13^, Lorena Orozco^12,28^, Colin N. A. Palmer^124^, James S. Pankow^125^, Anthony J. Payne^115^, Oluf B. Pedersen^62,126^, Catherine Pihoker^127^, Wendy S. Post^128^, Michael Preuss^20^, Bruce M. Psaty^24,129,130,131^, Asif Rasheed^54^, Alexander P. Reiner^132,133^, Cristina Revilla-Monsalve^134^, Stephen S. Rich^135^, Neil R. Robertson^101^, Jerome I. Rotter*^136^, Danish Saleheen^54,137^, Nicola Santoro^138,139^, Claudia Schurmann^20,140^, Laura J. Scott^1^, Mark Seielstad^141,142,143^, Yoon Shin Cho^144^, E. Shyong Tai^34,38,145^, Xueling Sim^1,98^, Robert Sladek^146,147,148^, Kerrin S. Small^149^, Xavier Soberón^12^, Kyong Soo Park^73,150,151^, Timothy D. Spector^149^, Konstantin Strauch^58,59,152^, Heather M. Stringham^1^, Tim M. Strom^107,108,109^, Claudia H. T. Tam^33^, Tanya M. Teslovich^1,13^, Farook Thameem^153^, Brian Tomlinson^154^, Jason M. Torres^115,155^, Russell P. Tracy^156^, Tiinamaija Tuomi^63,157,158,159^, Jaakko Tuomilehto^160,161,162,163,164^, Teresa Tusié-Luna^4,165^, Miriam S. Udler^8,35^, Rob M. van Dam^34,98,166^, Ramachandran S. Vasan^70,167^, Marijana Vujkovic^137^, Shuai Wang^47^, Ryan P. Welch^16^, Jennifer Wessel^168,169,170^, N. William Rayner^101,171^, James G. Wilson^172^, Daniel R. Witte^78,173,174^, Tien-Yin Wong^39,145,175^, Wing Yee So^154^, Mi Yeong Hwang^67^, Yik Ying Teo^176,177^, Philip Zeitler^25,43^

### Affiliations

1. Department of Biostatistics, Center for Statistical Genetics, University of Michigan, Ann Arbor, MI, USA

2. Regeneron Pharmaceuticals, Tarrytown, NY, USA

3. Instituto Nacional de Ciencias Médicas y Nutrición Salvador Zubirán, Mexico City, Mexico

4. Instituto Nacional de Ciencias Médicas y Nutrición, Mexico City, Mexico

5. Center for Genomic Medicine, Massachusetts General Hospital, Boston, MA, USA

6. Department of Biology, Massachusetts Institute of Technology, Cambridge, MA, USA

7. Department of Molecular Biology, Harvard Medical School, Boston, MA, USA

8. Programs in Metabolism and Medical and Population Genetics, Broad Institute of MIT and Harvard, Cambridge, MA, USA

9. Department of Medicine, Department of Genetics, Albert Einstein College of Medicine, New York, NY, USA

10. Departments of Medicine and Genetics, Albert Einstein College of Medicine, NY, USA

11. University of Haifa, Faculty of Natural Science, Haifa, Israel

12. Instituto Nacional de Medicina Genómica, Mexico City, Mexico

13. Regeneron Genetics Center, Regeneron Pharmaceuticals, Tarrytown, NY, USA

14. Department of Human Genetics, University of Chicago, Chicago, IL, USA

15. Department of Human Genetics and South Texas Diabetes and Obesity Institute, University of Texas, Rio Grande Valley, Brownsville, TX, USA

16. Department of Biostatistics and Center for Statistical Genetics, University of Michigan, Ann Arbor, MI, USA

17. Human Genetics Center, Department of Epidemiology, Human Genetics, and Environmental Sciences, School of Public Health, The University of Texas Health Science Center at Houston, Houston, Texas, USA

18. Human Genome Sequencing Center, Baylor College of Medicine, Houston, TX, USA

19. National Human Genome Research Institute, National Institutes of Health, Bethesda, MD, USA

20. Charles R. Bronfman Institute of Personalized Medicine, Mount Sinai School of Medicine, New York, NY, USA

21. The Charles Bronfman Institute for Personalized Medicine, Icahn School of Medicine at Mount Sinai, New York, NY, USA

22. Department of Biochemistry, Wake Forest School of Medicine, Winston-Salem, NC, USA

23. Wake Forest University, Winston-Salem, NC, USA

24. Cardiovascular Health Research Unit, University of Washington, Seattle, WA, USA

25. Biostatistics Center, George Washington University, Washington, DC, USA

26. Geisinger Health System, Danville, PA, USA

27. Geisinger, Danville, PA, USA

28. Instituto Nacional de Medicina Genómica, Mexico City, Mexico

29. Department of Cardiology, Ealing Hospital NHS Trust, Southall, Middlesex, UK

30. Department of Epidemiology and Biostatistics, Imperial College London, London, UK

31. Ealing Hospital National Health Service (NHS) Trust, Middlesex, UK

32. Imperial College Healthcare NHS Trust, London, UK

33. Li Ka Shing Institute of Health Sciences, Chinese University of Hong Kong, Hong Kong, China

34. Department of Medicine, Yong Loo Lin School of Medicine, National University of Singapore, Singapore

35. Diabetes Research Center (Diabetes Unit), Department of Medicine, Massachusetts General Hospital, Boston, MA, USA

36. Office of Clinical Sciences, Duke-NUS Graduate Medical School, National University of Singapore, Singapore

37. Ophthalmology and Visual Sciences Academic Clinical Program (Eye ACP), Duke-NUS Medical School, Singapore

38. Saw Swee Hock School of Public Health, National University of Singapore, Singapore

39. Singapore Eye Research Institute, Singapore National Eye Centre, Singapore

40. Department of Medicine, University of Mississippi Medical Center, Jackson, MS, USA

41. Broad Institute of MIT and Harvard, Cambridge, MA, USA

42. Vanderbilt Genetics Institute, Vanderbilt University, Nashville, TN, USA

43. Children’s Hospital Colorado, Aurora, CO, USA

44. Department of Epidemiology, Colorado School of Public Health, Aurora, CO, USA

45. Department of Medicine, University of Texas Health San Antonio, San Antonio, TX, USA

46. Cincinnati Children’s Hospital Medical Center, Cincinnati, OH, USA

47. Department of Biostatistics, Boston University School of Public Health, Boston, MA, USA

48. Centro de Estudios en Diabetes, Mexico City, Mexico

49. Departments of Medicine and Human Genetics, University of Chicago, Chicago, IL, USA

50. Department of Pediatrics, Harvard Medical School, Boston, MA, USA

51. Division of Genetics and Genomics, Boston Children’s Hospital, Boston, MA, USA

52. Department of Medicine, Harvard Medical School, Boston, MA, USA

53. Department of Medicine and Epidemiology, University of Washington, Seattle, WA, USA

54. Center for Non-Communicable Diseases, Karachi, Pakistan

55. Institute for Biomedicine, Eurac Research, Bolzano, Italy

30. 56. Genomics Platform, Broad Institute of Harvard and MIT, Cambridge, MA, USA

57. German Center for Diabetes Research (DZD e.V.), Neuherberg, Germany

58. Institute of Genetic Epidemiology, Helmholtz Zentrum München, Neuherberg, Germany

59. Research Unit of Molecular Epidemiology, Institute of Epidemiology, Helmholtz Zentrum München, German Research Center for Environmental Health, Neuherberg, Germany

60. Department of Endocrinology and Metabolism, Hadassah Medical Center and Faculty of Medicine, Hebrew University of Jerusalem, Israel

61. Unidad de Diabetes y Riesgo Cardiovascular, Instituto Nacional de Salud Pública, Cuernavaca, Morelos, Mexico

62. The Novo Nordisk Foundation Center for Basic Metabolic Research, Faculty of Health and Medical Sciences, University of Copenhagen, Copenhagen, Denmark

63. Department of Clinical Sciences, Diabetes and Endocrinology, Lund University Diabetes Centre, Malmö, Sweden

64. Institute for Molecular Genetics Finland, University of Helsinki, Helsinki, Finland

65. Department of Laboratory Medicine and Pathology, University of Minnesota, Minneapolis, MN, USA

66. Department of Preventive Medicine, Keck School of Medicine, University of Southern California, Los Angeles, CA, USA

67. Division of Genome Science, Department of Precision Medicine, National Institute of Health, Chungcheongbuk-do, Republic of Korea

68. Faculty of Health Sciences, University of Southern Denmark, Odense, Denmark

69. Boston University, Boston, MA, USA

70. National Heart, Lung, and Blood Institute Framingham Heart Study, Framingham, MA, USA

71. Department of Neurology, Boston University School of Medicine, Boston, MA, USA

72. Cardiovascular Health Research Unit and Department of Epidemiology, University of Washington, Seattle, WA, USA

73. Department of Internal Medicine, Seoul National University Hospital, Seoul, Republic of Korea

74. Department of Pathology, University of Michigan, Ann Arbor, MI, USA

75. Greenland Centre for Health Research, University of Greenland, Nuuk, Greenland

76. National Institute of Public Health, University of Southern Denmark, Odense, Denmark

77. Steno Diabetes Center Copenhagen, Gentofte, Denmark

78. Steno Diabetes Center, Gentofte, Denmark

79. Department of Public Health Solutions, National Institute for Health and Welfare, Helsinki, Finland

80. Minerva Foundation Institute for Medical Research, Helsinki, Finland

81. University of Helsinki and Department of Medicine, Helsinki University Central Hospital, Helsinki, Finland

82. National Heart and Lung Institute (NHLI), Imperial College London, Hammersmith Hospital, London, UK

83. National Heart and Lung Institute, Cardiovascular Sciences, Hammersmith Campus, Imperial College London, London, UK

84. Department of Medicin, Kuopio University Hospital, Kuopio, Finland

85. Institute of Clinical Medicine, Internal Medicine, University of Eastern Finland and Kuopio University Hospital, Kuopio, Finland

86. Institute of Clinical Medicine, Internal Medicine, University of Eastern Finland, Kuopio, Finland

87. Department of Medicine, University of Eastern Finland, Kuopio Campus and Kuopio University Hospital, Kuopio, Finland

88. Department of Genetics, University of North Carolina, Chapel Hill, NC, USA

89. Department of Medicine, University of Colorado Denver, Anschutz Medical Campus, Aurora, CO, USA

90. University of North Carolina Chapel Hill, Chapel Hill, NC, USA

91. Center for Genome Science, Korea National Institute of Health, Osong Health Technology Administration Complex, Chungcheongbuk-do, South Korea

92. Department of Business Data Convergence, Chungbuk National University, Gyeonggi-do, Republic of Korea

93. Center for Clinical Research and Prevention, Bispebjerg and Frederiksberg Hospital, Copenhagen, Denmark

94. Department of Clinical Experimental Research, Rigshospitalet, Copenhagen, Denmark

95. Department of Clinical Medicine, Faculty of Health and Medical Sciences, University of Copenhagen, Copenhagen, Denmark

96. Research Centre for Prevention and Health, Glostrup University Hospital, Glostrup, Denmark

97. Genome Institute of Singapore, Agency for Science Technology and Research, Singapore

98. Saw Swee Hock School of Public Health, National University of Singapore, Singapore

99. The Mindich Child Health and Development Institute, Icahn School of Medicine at Mount Sinai, New York, NY, USA

100. University of Bergen, Bergen, Norway

101. Wellcome Trust Centre for Human Genetics, University of Oxford, Oxford, UK

102. Clinical and Translational Epidemiology Unit, Massachusetts General Hospital, Harvard University, Cambridge, MA, USA

103. Massachusetts General Hospital, Boston, MA, USA

104. University of North Carolina, Chapel Hill, NC, USA

105. General Medicine Division, Massachusetts General Hospital, Boston, MA, USA

106. Deutsches Forschungszentrum für Herz-Kreislauferkrankungen (DZHK), Partner Site Munich Heart Alliance, Munich, Germany

107. Human Genetics, Helmholtz Zentrum München, Neuherberg, Germany

108. Institute of Human Genetics, Helmholtz Zentrum München, German Research Center for Environmental Health, Neuherberg, Germany

109. Institute of Human Genetics, Technische Universität München, Munich, Germany

110. Center for Human Genetic Research and Diabetes Research Center (Diabetes Unit), Massachusetts General Hospital, Boston, MA, USA

111. Joint BSC-CRG-IRB Research Program in Computational Biology, Barcelona Supercomputing Center, Barcelona, Spain

112. Department of Genetics, University of North Carolina Chapel Hill, Chapel Hill, NC, USA

113. Department of Biostatistics, University of Liverpool, Liverpool, UK

114. Department of Genetic Medicine, Manchester Academic Health Sciences Centre, Manchester, UK

115. Wellcome Centre for Human Genetics, Nuffield Department of Medicine, University of Oxford, Oxford, UK

116. Clinical Research Centre, Centre for Molecular Medicine, Ninewells Hospital and Medical School, Dundee, UK

117. Center for Genomics and Personalized Medicine Research, Center for Diabetes Research, Department of Biochemistry, Department of Internal Medicine, Wake Forest School of Medicine, Winston-Salem, NC, USA

118. Department of Clinical Sciences, Medicine, Lund University, Malmö, Sweden

119. Cardiology Division, Massachusetts General Hospital, Boston, MA, USA

120. Brigham and Women’s Hospital, Boston, MA, USA

121. Intramural Administration Management Branch, National Heart Lung and Blood Institute, NIH, Framingham, MA, USA

122. National Heart, Lung, and Blood Institute, Bethesda, MD, USA

123. Section of Cardiology, Department of Medicine, VA Boston Healthcare, Boston, MA, USA

124. Pat Macpherson Centre for Pharmacogenetics and Pharmacogenomics, Medical Research Institute, Ninewells Hospital and Medical School, Dundee, UK

125. Division of Epidemiology and Community Health, School of Public Health, University of Minnesota, Minneapolis, MN, USA

126. Hagedorn Research Institute, Gentofte, Denmark

127. Seattle Children’s Hospital, Seattle, WA, USA

128. Division of Cardiology, Department of Medicine, Johns Hopkins University, Baltimore, MD, USA

129. Group Health Research Institute, Group Health Cooperative, Seattle, WA, USA

130. Group Health Research Institute, Seattle, WA, USA

131. Kaiser Permanente Washington Health Research Institute, Seattle, WA, USA

132. Fred Hutchinson Cancer Research Center, Seattle, WA, USA

133. University of Washington, Seattle, WA, USA

134. Instituto Mexicano del Seguro Social SXXI, Mexico City, Mexico

135. Center for Public Health Genomics, University of Virginia School of Medicine, Charlottesville, VA, USA

136. The Institute for Translational Genomics and Population Sciences, Department of Pediatrics, The Lundquist Institute for Biomedical Innovation at Harbor-UCLA Medical Center, Torrance, CA 90502 USA

137. Department of Biostatistics and Epidemiology, University of Pennsylvania, Philadelphia, PA, USA

138. Department of Pediatrics, Yale University, New Haven, CT, USA

139. Yale School of Medicine, New Haven, CT, USA

140. The Genetics of Obesity and Related Metabolic Traits Program, Icahn School of Medicine at Mount Sinai, New York, NY, USA

141. Blood Systems Research Institute, San Francisco, CA, USA

142. Department of Laboratory Medicine and Institute for Human Genetics, University of California, San Francisco, San Francisco, CA, USA

143. University of California San Francisco, San Francisco, CA, USA

144. Department of Biomedical Science, Hallym University, Gangwon-do, South Korea

145. Duke-NUS Medical School, Singapore

146. Department of Medicine, Royal Victoria Hospital, Montréal, Québec, Canada

147. Division of Endocrinology and Metabolism, Department of Medicine, McGill University, Montréal, Québec, Canada

148. McGill University and Génome Québec Innovation Centre, Montreal, Quebec, Canada

149. Department of Twin Research and Genetic Epidemiology, King’s College London, London, UK

150. Department of Internal Medicine, Seoul National University College of Medicine, Seoul, Republic of Korea

151. Department of Molecular Medicine and Biopharmaceutical Sciences, Graduate School of Convergence Science and Technology, Seoul National University, Seoul, South Korea

152. Institute of Medical Informatics, Biometry and Epidemiology, Chair of Genetic Epidemiology, Ludwig-Maximilians-Universität, Neuherberg, Germany

153. Department of Biochemistry, Faculty of Medicine, Health Science Center, Kuwait University, Safat, Kuwait

154. Department of Medicine and Therapeutics, Chinese University of Hong Kong, Hong Kong, China

155. Department of Medicine, University of Chicago, Chicago, IL, USA

156. Department of Biochemistry, Robert Larner M.D. College of Medicine, University of Vermont, Burlington, VT, USA

157. Department of Endocrinology, Abdominal Centre, Helsinki University Hospital, Helsinki, Finland

158. Folkhälsan Research Centre, Helsinki, Finland

159. Research Programs Unit, Diabetes and Obesity, University of Helsinki, Helsinki, Finland

160. Center for Vascular Prevention, Danube University Krems, Krems, Austria

161. Department of Public Health, University of Helsinki, Helsinki, Finland

162. Diabetes Prevention Unit, National Institute for Health and Welfare, Helsinki, Finland

163. Diabetes Research Group, King Abdulaziz University, Jeddah, Saudi Arabia

164. Instituto de Investigacion Sanitaria del Hospital Universario LaPaz (IdiPAZ), University Hospital La-Paz, Autonomous University of Madrid, Madrid, Spain

165. Instituto de Investigaciones Biomédicas, Departamento de Medicina Genómica y ToxicologiÌĄa, Universidad Nacional Autónoma de México, Mexico City, Mexico

166. Department of Nutrition, Harvard School of Public Health, Boston, MA, USA

167. Preventive Medicine and Epidemiology, Medicine, Boston University School of Medicine, Boston, MA, USA

168. Department of Epidemiology, Fairbanks School of Public Health, Indianapolis, IN, USA

169. Department of Medicine, Indiana University School of Medicine, Indianapolis, IN, USA

170. Department of Medicine, School of Medicine, Indiana University, Indianapolis, IN, USA

171. Department of Human Genetics, Wellcome Trust Sanger Institute, Hinxton, Cambridgeshire, UK

172. Department of Physiology and Biophysics, University of Mississippi Medical Center, Jackson, MS, USA

173. Danish Diabetes Academy, Odense, Denmark

174. Department of Public Health, Aarhus University, Aarhus, Denmark

175. Department of Ophthalmology, Yong Loo Lin School of Medicine, National University of Singapore, National University Health System, Singapore

176. Department of Epidemiology and Public Health, National University of Singapore, Singapore

177. Genome Institute of Singapore, Agency for Science, Technology and Research, Singapore

* supported in part by the National Institute of Diabetes and Digestive and Kidney Disease Diabetes Research Center (DRC) grant DK063491 to the Southern California Diabetes Endocrinology Research Center.

